# Smoking and COVID-19: A two-sample Mendelian randomization study

**DOI:** 10.1101/2021.03.31.21254730

**Authors:** Bing-Kun Zheng, Na Li

**Affiliations:** Neonatal Intensive Care Unit, First Affiliated Hospital of Zhengzhou University, Zhengzhou, China

## Abstract

Evidence from observational studies suggested that smokers are at increased risk of coronavirus disease 2019 (COVID-19). We aimed to assess the causal effect of smoking on risk for COVID-19 susceptibility and severity using two-sample Mendelian randomization method. Smoking-associated variants were selected as instrument variables from two largest genetic studies. The latest summary data of COVID-19 that shared on Jan 18, 2021 by the COVID-19 Host Genetics Initiative was used. The present Mendelian randomization study provided genetic evidence that smoking was a causal risk factor for COVID-19 susceptibility and severity. In addition, there may be a dose-effect relationship between smoking and COVID-19 severity.

## Background

Evidence from observational studies suggested that smokers are at increased risk of coronavirus disease 2019 (COVID-19) (1, 2). Two-sample Mendelian randomization (MR) study could be used to assess the effect of smoking on risk of COVID-19 because it could assess causality and could limit the bias caused by confounders (3). We aimed to assess the causal effect of smoking on risk for COVID-19 susceptibility and severity using two-sample MR method.

## Methods

Four following measures of smoking were chosen as exposures: smoking initiation: defined as ever being a regular smoker; lifetime smoking: a continuous variable that accounting for smoking status, age at smoking initiation and cessation, cigarettes per day, and a simulated half-life constant effected on health outcomes; cigarettes per day: defined as the average number of cigarettes smoked per day; and smoking cessation: defined as current smoking vs. former smoking. Independent and genome-wide significant single nucleotide polymorphisms (SNPs) for these measures were selected from two large genetic studies (Figure 1) (4, 5).

**Figure 1.**
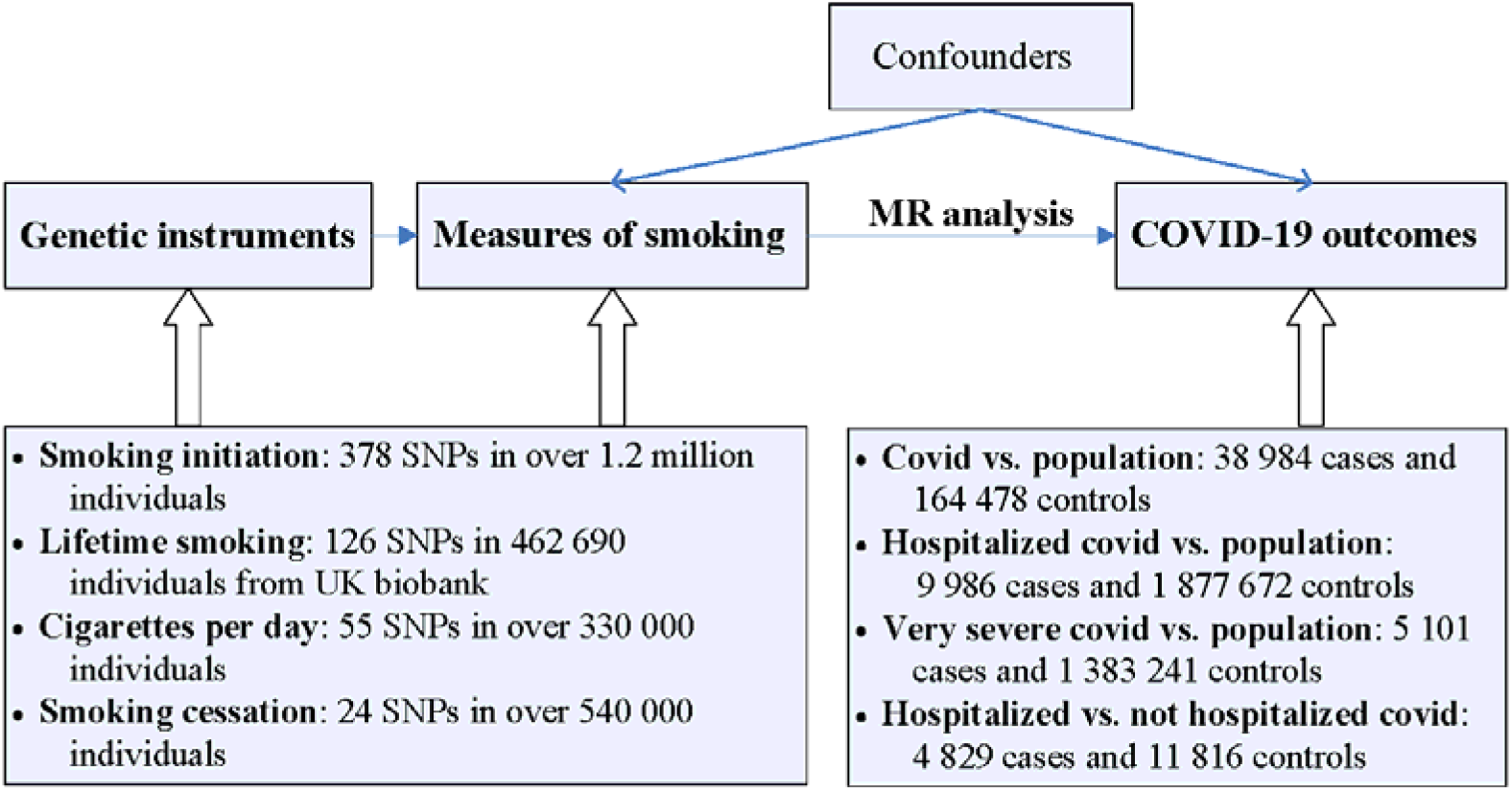
Overview of the study design. All individuals were of European ancestry. For exposure of lifetime smoking, we used summary data of COVID-19 that excluding UK biobank data.

We used the latest summary data of COVID-19 that shared on Jan 18, 2021 by the COVID-19 Host Genetics Initiative (Figure 1) (6).

Two-sample MR analyses were performed using the TwoSampleMR package in R 3.6.0.(3) We chose random-effects inverse variance weighted method as the main MR method. Several other MR methods were chosen to perform sensitivity analysis. Additional sensitivity analysis was performed by excluding SNPs associated (p < 5×10^−8^) with body mass index (BMI) because previous MR study showed BMI was the only one cardiometabolic factor that associated with COVID-19 (7). P value of MR analysis small than 0.003 (0.05/16) was considered as statistically significant. P value of MR analysis small than 0.05 was considered as nominally significant.

## Results

Main MR analyses showed smoking initiation was significantly or nominally associated with outcomes of COVID-19 cases vs. population controls, hospitalized COVID-19 cases vs. population controls, and very severe respiratory confirmed COVID-19 cases vs. population controls. The odds ratios (ORs) and 95% confidence intervals (CIs) were 1.15 (1.07–2.13, p = 1.27×10^−4^), 1.33 (1.16–1.51, p = 2.35×10^−5^), and 1.29 (1.07–1.56, p = 0.009) respectively (Figure 2). The ORs were 1.14 (p = 8.84×10^−4^), 1.30 (3.71×10^−4^), and 1.33 (p = 0.008) after excluding BMI-associated SNPs.

**Figure 2.**
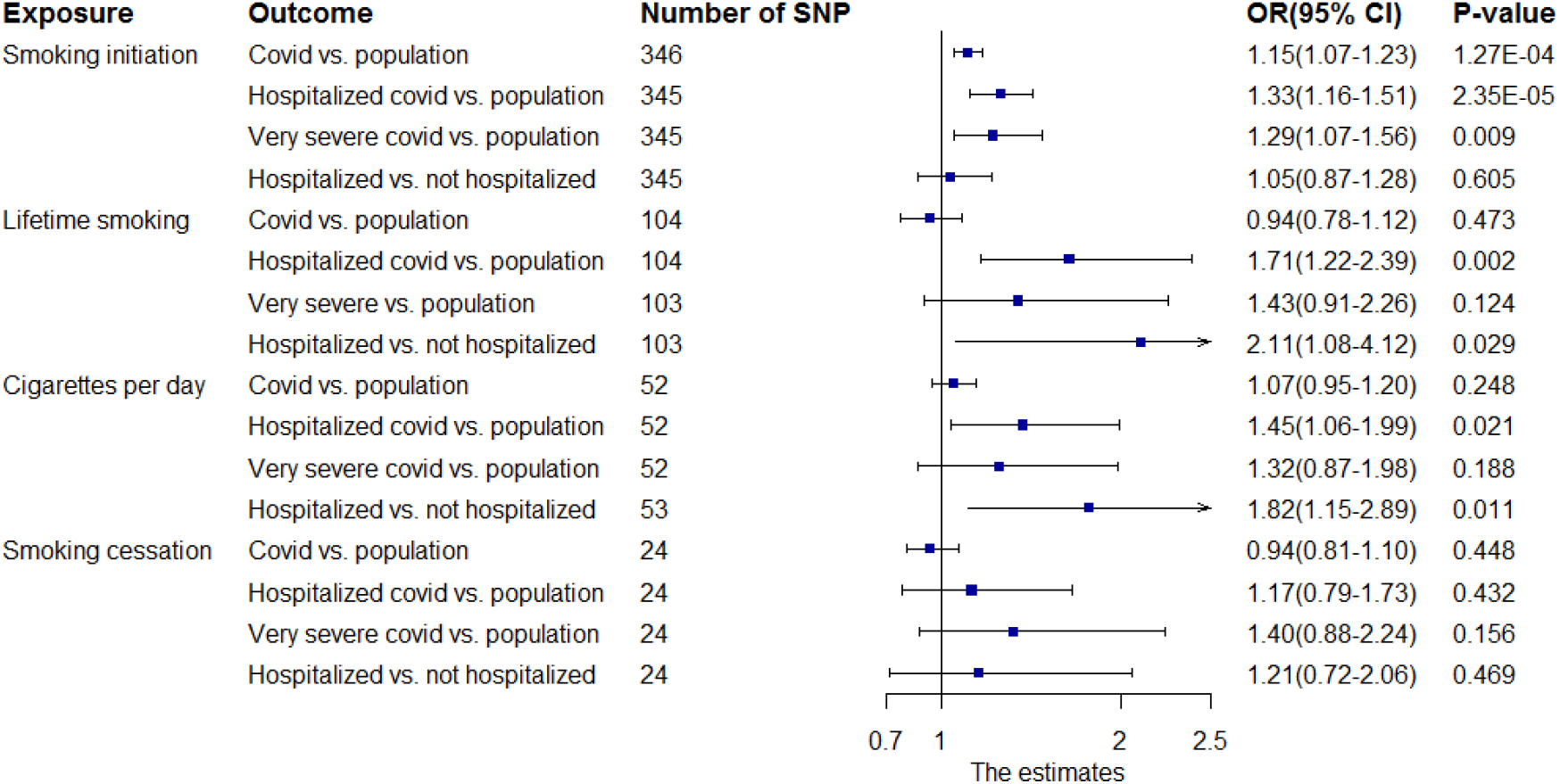
Two-sample MR analyses of the effect of smoking on COVID-19 susceptibility and severity. All of the results are based on random-effects inverse variance weighted method. A proxy SNP (R^2^>0.8) was chosen (if available) if an exposure-associated SNP was not found for outcome.

Main MR analyses showed lifetime smoking was significantly associated with hospitalized COVID-19 cases vs. population controls (OR = 1.71, 95% CI = 1.22–2.39, p = 0.002) and nominally associated with hospitalized COVID-19 cases vs. not hospitalized COVID-19 controls (OR = 2.11, 95% CI = 1.08–4.12, p = 0.029) (Figure 2). The ORs were 1.66 (p = 0.007) and 2.24 (p = 0.031) after excluding BMI-associated SNPs.

Main MR analyses showed cigarettes per day was nominally associated with hospitalized COVID-19 cases vs. population controls (OR = 1.45, 95% CI = 1.06–1.99, p = 0.021) and hospitalized COVID-19 cases vs. not hospitalized COVID-19 controls (OR = 1.82, 95% CI = 1.15–2.89, p = 0.011) (Figure 2). The Ors were 1.41 (p = 0.048) and 1.78 (p = 0.023) after excluding BMI-associated SNPs.

Main MR analyses showed no significant effect of smoking cessation on any of the four COVID-19 outcomes (Figure 2).

Although the significant results were not confirmed in some sensitivity analyses using other MR methods, no evidence of unbalanced pleiotropy was found by MR Egger regression (p > 0.097).

Supplemental Figures show the SNP effects on exposures and outcomes across multiple MR methods.

## Discussion

We could not assess the effect of current smoking and former smoking separately because the data was not available. Meta-analyses suggested that current smokers had higher odds compared with non-current smokers but lower odds compared with former smokers for COVID-19 susceptibility and adverse outcome (1, 2). This finding was not supported by the present study because the present study showed current smoking was not associated with decreased risk of COVID-19 susceptibility and severity compared with former smoking. However, the statistical power is relatively small for exposure of smoking cessation in the present study. In addition, time since cessation may also associated with COVID-19 risk (1).

## Conclusion

The present MR study provided genetic evidence that smoking was a causal risk factor for COVID-19 susceptibility and severity. In addition, there may be a dose-effect relationship between smoking and COVID-19 severity.

## Data Availability

All analyses were based on publicly available summary data from published genetics studies.

## Financial support information

None.

## Acknowledgments

We thank the COVID-19 Host Genetics Initiative and all other contributors for making summary statistics publicly accessible.

