## Supplemental Figures for "Smoking and COVID-19: A two-sample Mendelian randomization study"

**Table of Contents**

**Supplemental Figure I** Scatter plots of the SNP effects on the exposure, smoking initiation, and the outcome, COVID-19 cases vs. population controls across multiple MR methods.

**Supplemental Figure II** Scatter plots of the SNP effects on the exposure, smoking initiation, and the outcome, hospitalized COVID-19 cases vs. population controls across multiple MR methods.

**Supplemental Figure III** Scatter plots of the SNP effects on the exposure, smoking initiation, and the outcome, very severe respiratory confirmed COVID-19 cases vs. population controls across multiple MR methods.

**Supplemental Figure IV** Scatter plots of the SNP effects on the exposure, smoking initiation, and the outcome, hospitalized COVID-19 cases vs. not hospitalized COVID-19 controls across multiple MR methods.

**Supplemental Figure V** Scatter plots of the SNP effects on the exposure, lifetime smoking, and the outcome, COVID-19 cases vs. population controls across multiple MR methods.

**Supplemental Figure VI** Scatter plots of the SNP effects on the exposure, lifetime smoking, and the outcome, hospitalized COVID-19 cases vs. population controls across multiple MR methods.

**Supplemental Figure VII** Scatter plots of the SNP effects on the exposure, lifetime smoking, and the outcome, very severe respiratory confirmed COVID-19 cases vs. population controls across multiple MR methods.

**Supplemental Figure VIII** Scatter plots of the SNP effects on the exposure, lifetime smoking, and the outcome, hospitalized COVID-19 cases vs. not hospitalized COVID-19 controls across multiple MR methods.

**Supplemental Figure IX** Scatter plots of the SNP effects on the exposure, cigarettes per day, and the outcome, COVID-19 cases vs. population controls across multiple MR methods.

**Supplemental Figure X** Scatter plots of the SNP effects on the exposure, cigarettes per day, and the outcome, hospitalized COVID-19 cases vs. population controls across multiple MR methods.

**Supplemental Figure XI** Scatter plots of the SNP effects on the exposure, cigarettes per day, and the outcome, very severe respiratory confirmed COVID-19 cases vs. population controls across multiple MR methods.

**Supplemental Figure XII** Scatter plots of the SNP effects on the exposure, cigarettes per day, and the outcome, hospitalized COVID-19 cases vs. not hospitalized COVID-19 controls across multiple MR methods.

**Supplemental Figure XIII** Scatter plots of the SNP effects on the exposure, smoking cessation, and the outcome, COVID-19 cases vs. population controls across multiple MR methods.

**Supplemental Figure XIV** Scatter plots of the SNP effects on the exposure, smoking cessation, and the outcome, hospitalized COVID-19 cases vs. population controls across multiple MR methods.

**Supplemental Figure XV** Scatter plots of the SNP effects on the exposure, smoking cessation, and the outcome, very severe respiratory confirmed COVID-19 cases vs. population controls across multiple MR methods.

**Supplemental Figure XVI** Scatter plots of the SNP effects on the exposure, smoking cessation, and the outcome, hospitalized COVID-19 cases vs. not hospitalized COVID-19 controls across multiple MR methods.

**Supplemental Figure I** Scatter plots of the SNP effects on the exposure, smoking initiation, and the outcome, COVID-19 cases vs. population controls across multiple MR methods.

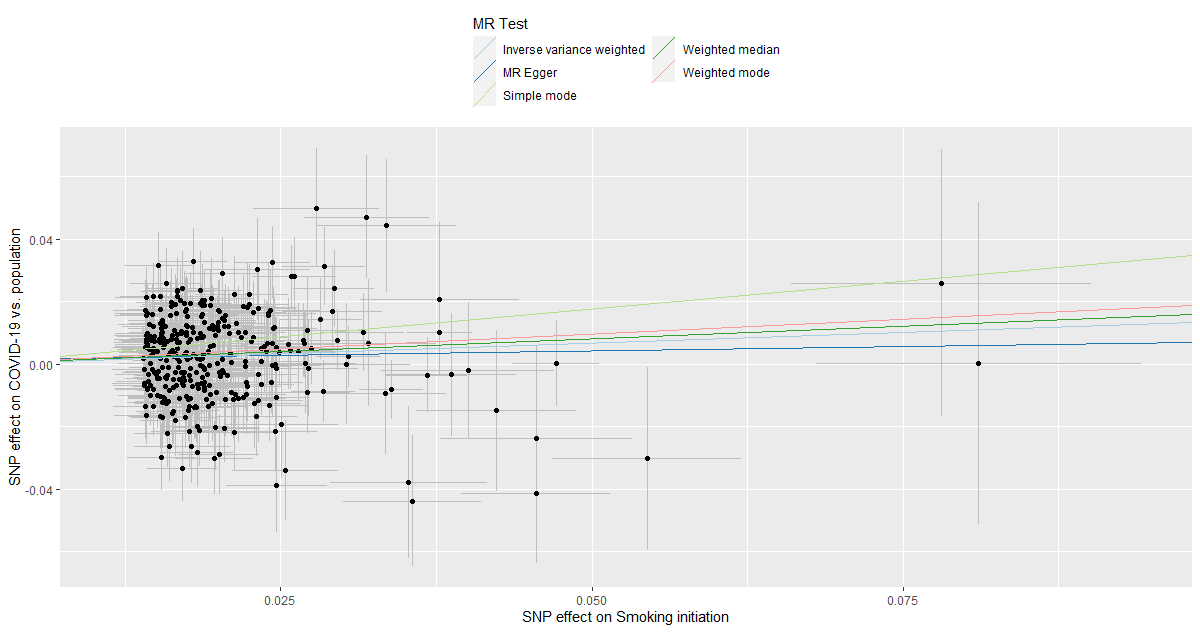

**Supplemental Figure II** Scatter plots of the SNP effects on the exposure, smoking initiation, and the outcome, hospitalized COVID-19 cases vs. population controls across multiple MR methods.

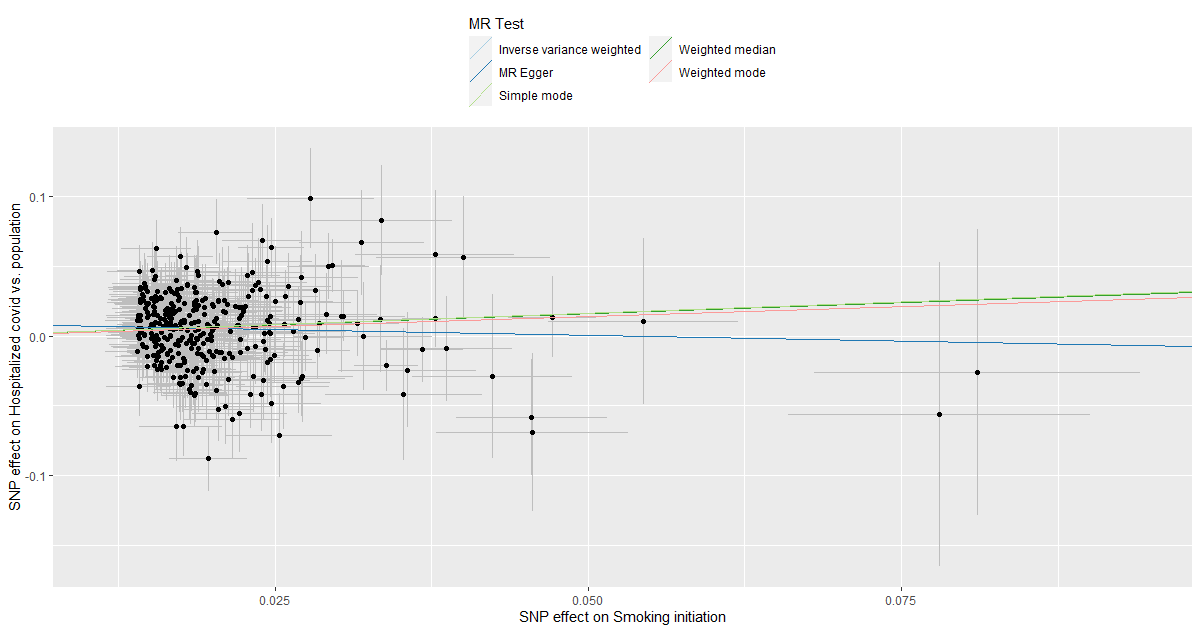

**Supplemental Figure III** Scatter plots of the SNP effects on the exposure, smoking initiation, and the outcome, very severe respiratory confirmed COVID-19 cases vs. population controls across multiple MR methods.

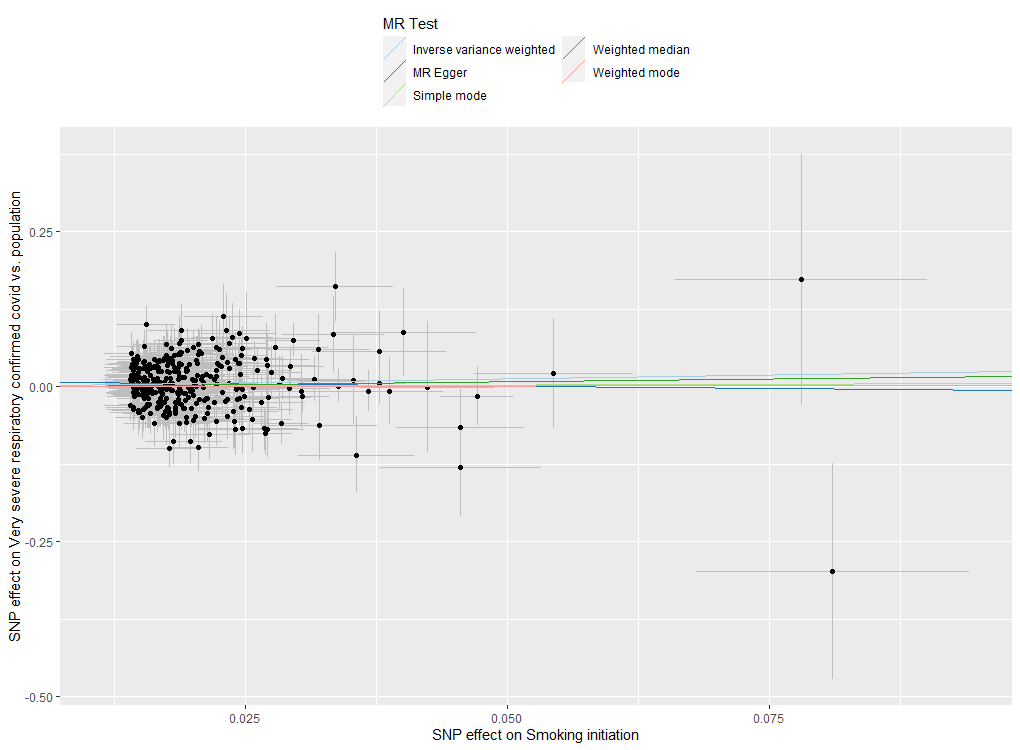

**Supplemental Figure IV** Scatter plots of the SNP effects on the exposure, smoking initiation, and the outcome, hospitalized COVID-19 cases vs. not hospitalized COVID-19 controls across multiple MR methods.

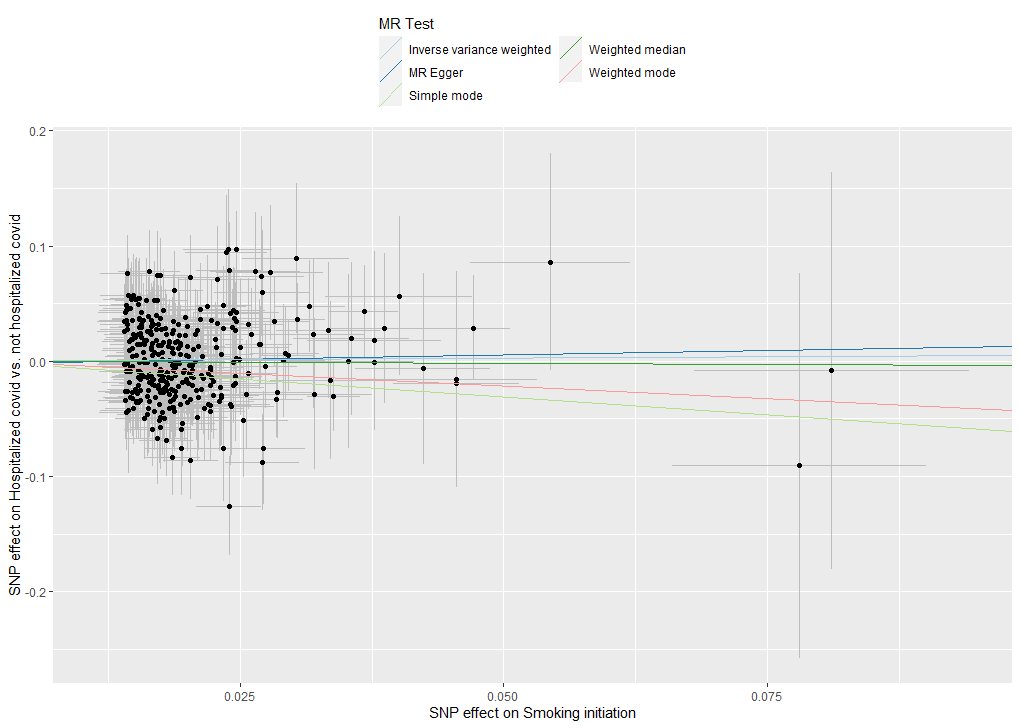

**Supplemental Figure V** Scatter plots of the SNP effects on the exposure, lifetime smoking, and the outcome, COVID-19 cases vs. population controls across multiple MR methods.

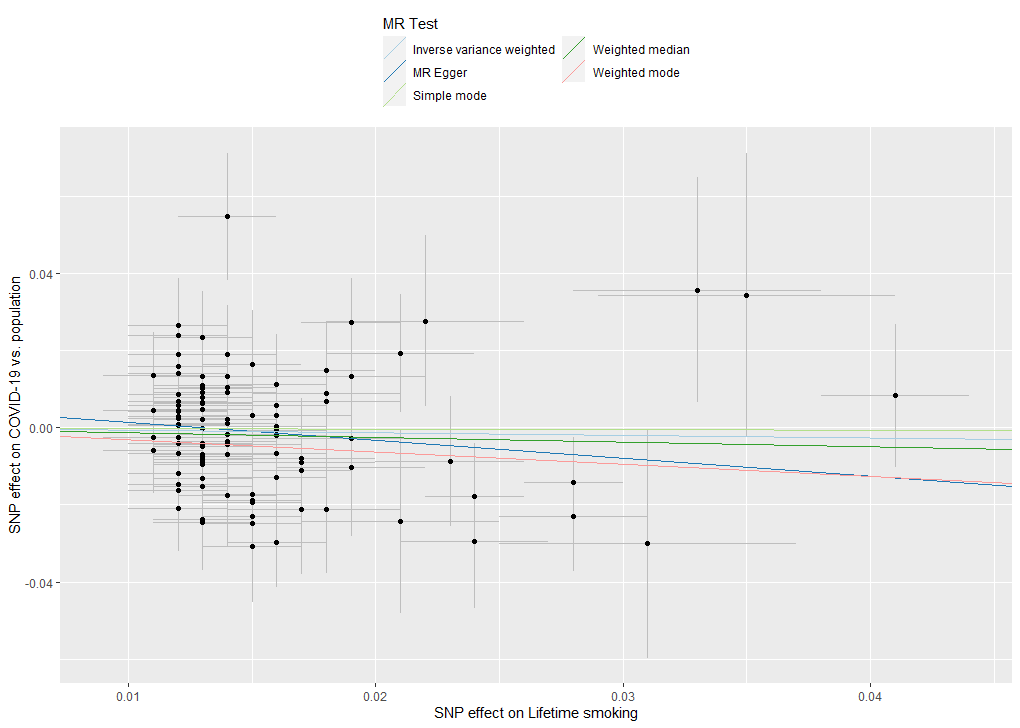

**Supplemental Figure VI** Scatter plots of the SNP effects on the exposure, lifetime smoking, and the outcome, hospitalized COVID-19 cases vs. population controls across multiple MR methods.

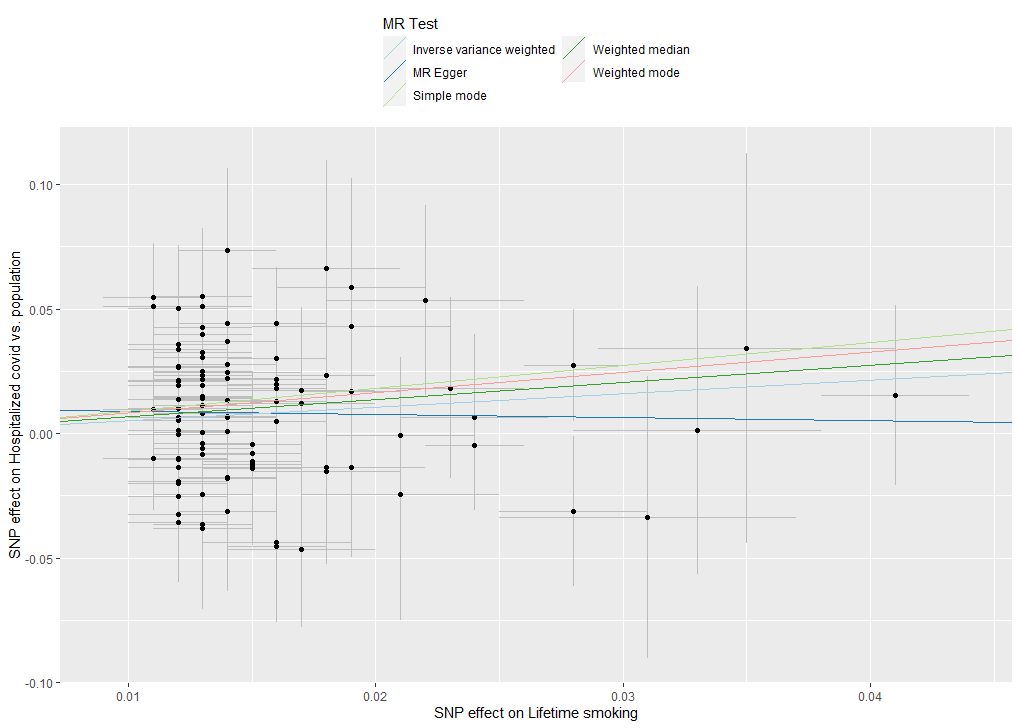

**Supplemental Figure VII** Scatter plots of the SNP effects on the exposure, lifetime smoking, and the outcome, very severe respiratory confirmed COVID-19 cases vs. population controls across multiple MR methods.

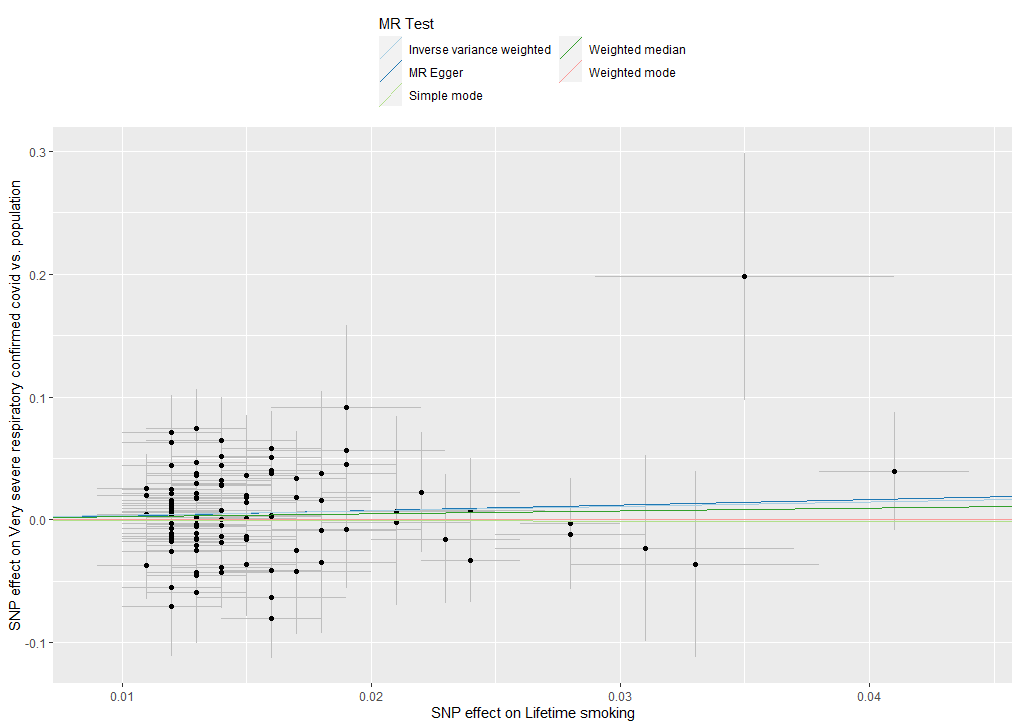

**Supplemental Figure VIII** Scatter plots of the SNP effects on the exposure, lifetime smoking, and the outcome, hospitalized COVID-19 cases vs. not hospitalized COVID-19 controls across multiple MR methods.

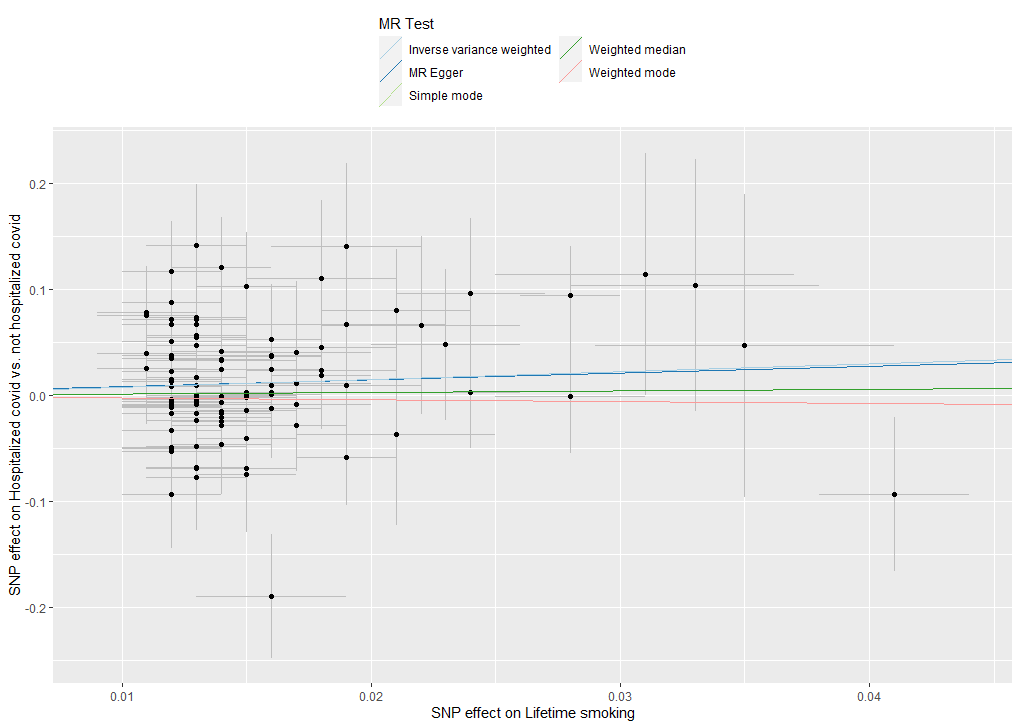

**Supplemental Figure IX** Scatter plots of the SNP effects on the exposure, cigarettes per day, and the outcome, COVID-19 cases vs. population controls across multiple MR methods.

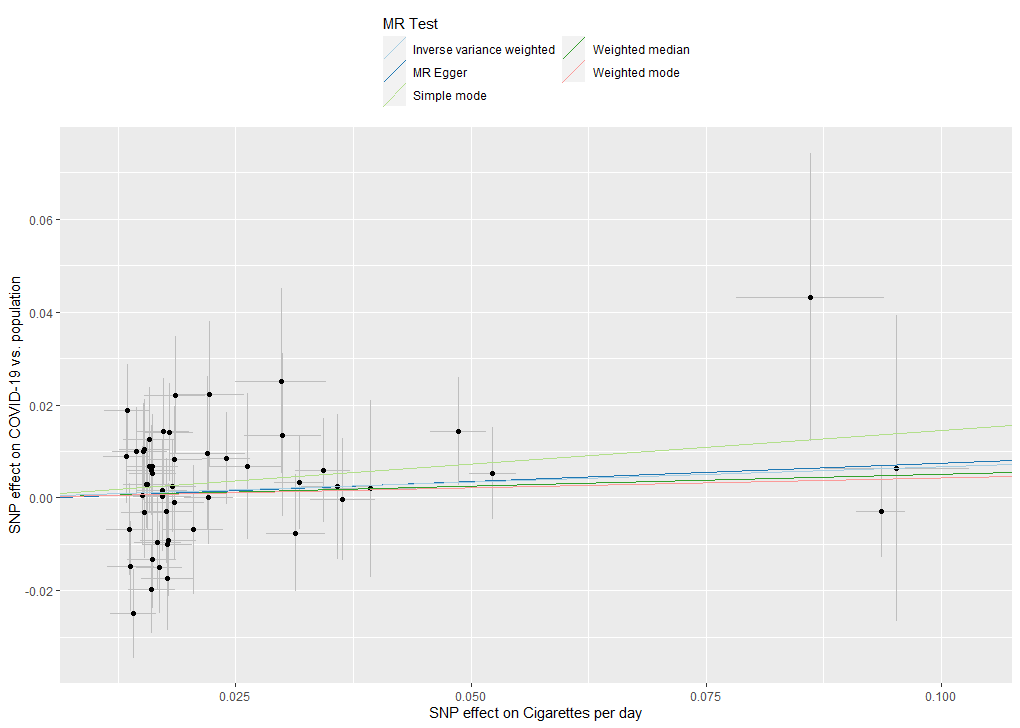

**Supplemental Figure X** Scatter plots of the SNP effects on the exposure, cigarettes per day, and the outcome, hospitalized COVID-19 cases vs. population controls across multiple MR methods.

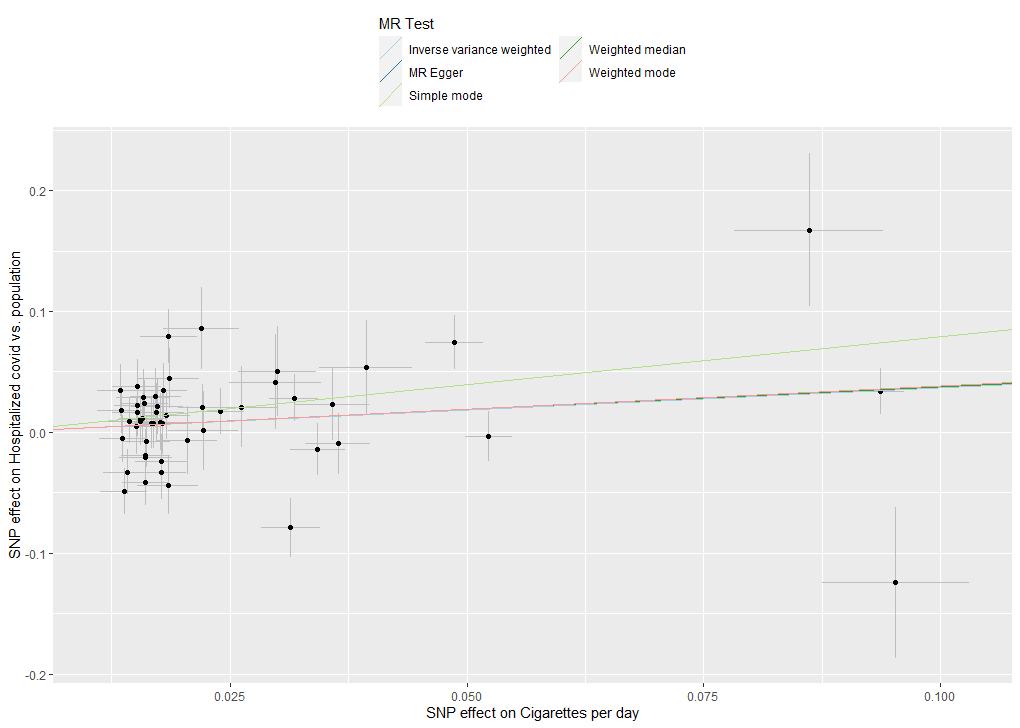

**Supplemental Figure XI** Scatter plots of the SNP effects on the exposure, cigarettes per day, and the outcome, very severe respiratory confirmed COVID-19 cases vs. population controls across multiple MR methods.

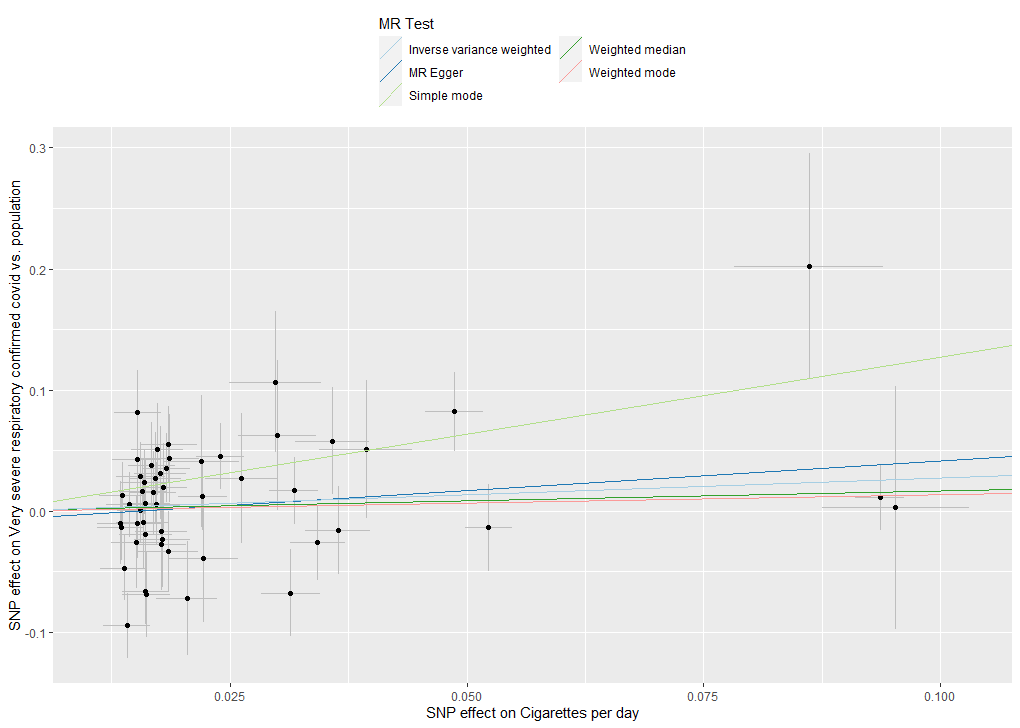

**Supplemental Figure XII** Scatter plots of the SNP effects on the exposure, cigarettes per day, and the outcome, hospitalized COVID-19 cases vs. not hospitalized COVID-19 controls across multiple MR methods.

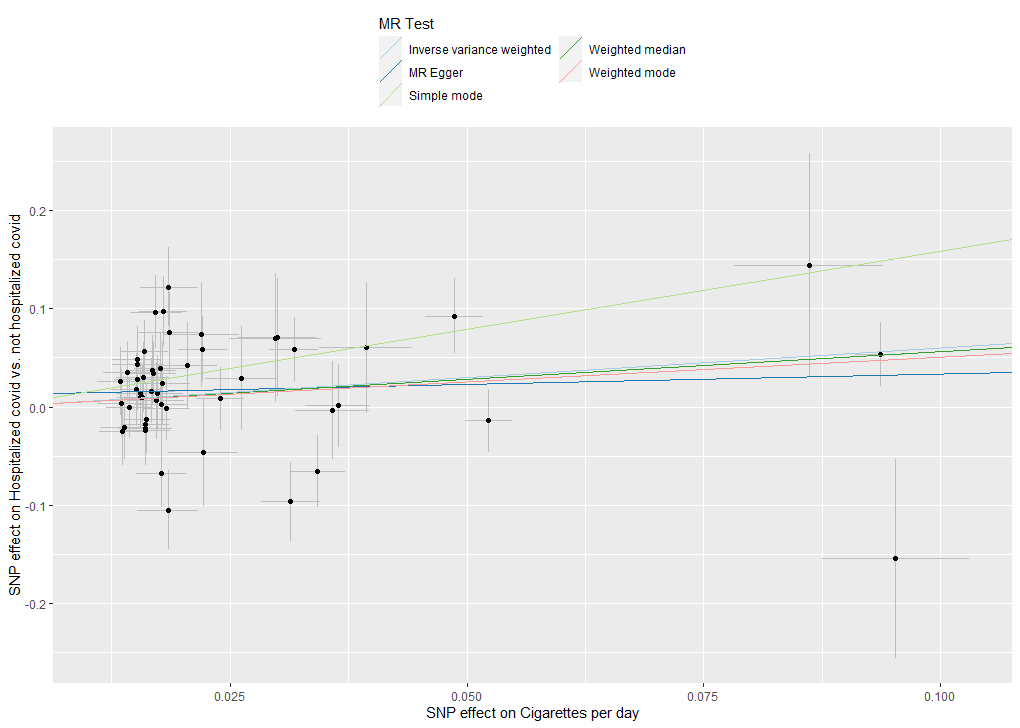

**Supplemental Figure XIII** Scatter plots of the SNP effects on the exposure, smoking cessation, and the outcome, COVID-19 cases vs. population controls across multiple MR methods.

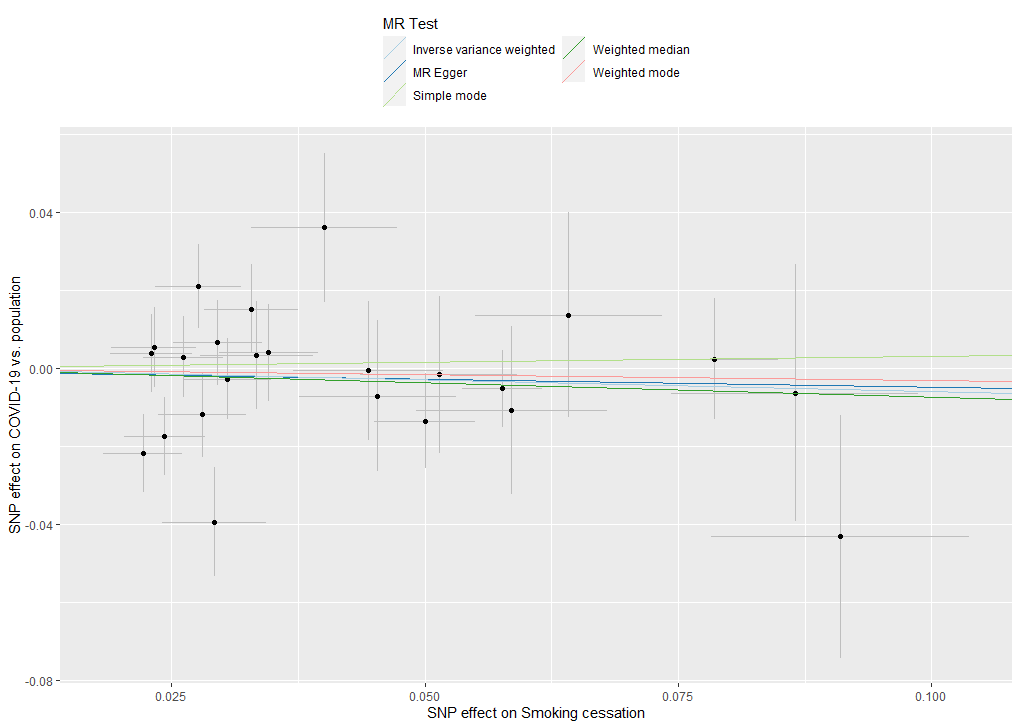

**Supplemental Figure XIV** Scatter plots of the SNP effects on the exposure, smoking cessation, and the outcome, hospitalized COVID-19 cases vs. population controls across multiple MR methods.

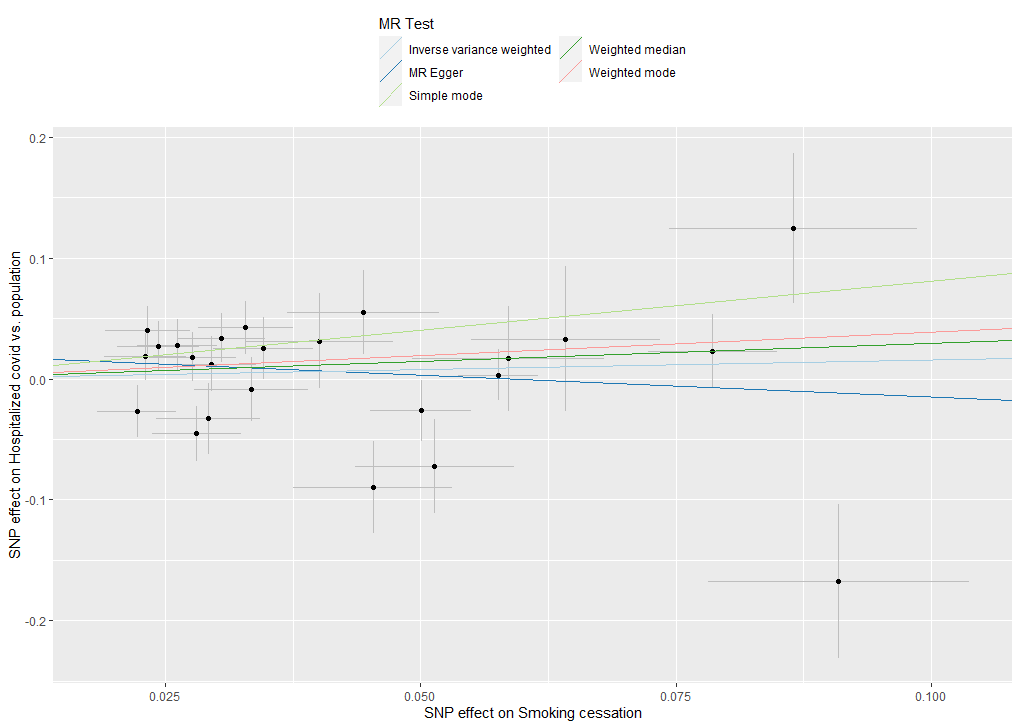

**Supplemental Figure XV** Scatter plots of the SNP effects on the exposure, smoking cessation, and the outcome, very severe respiratory confirmed COVID-19 cases vs. population controls across multiple MR methods.

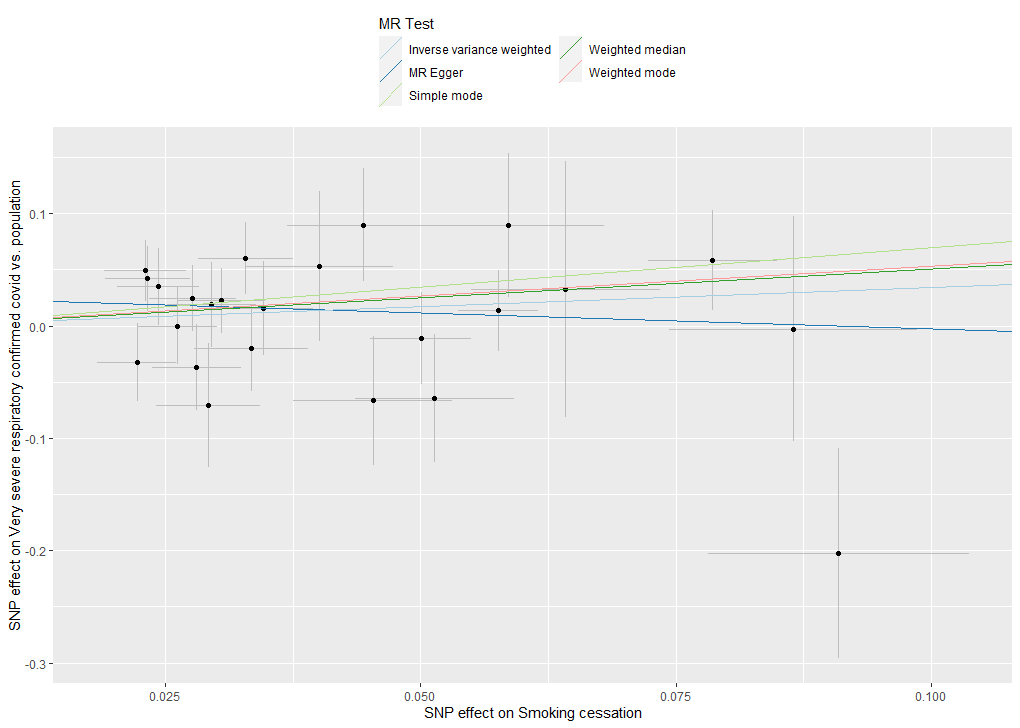

**Supplemental Figure XVI** Scatter plots of the SNP effects on the exposure, smoking cessation, and the outcome, hospitalized COVID-19 cases vs. not hospitalized COVID-19 controls across multiple MR methods.

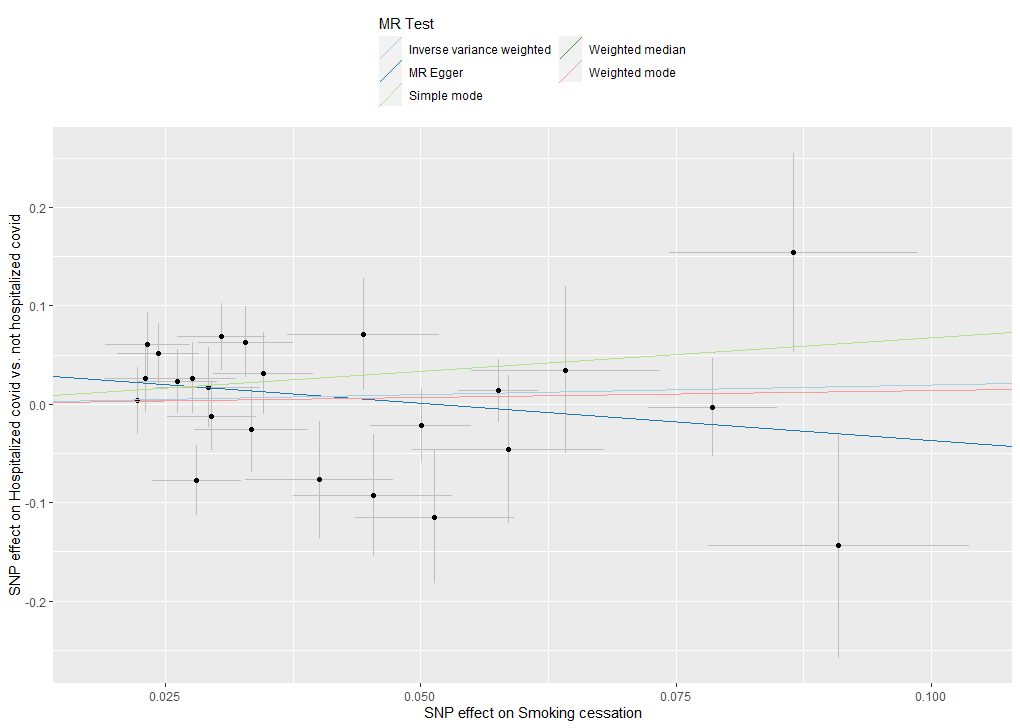
